# Multi-trait genome-wide analysis identified novel risk loci and candidate drugs for heart failure

**DOI:** 10.1101/2024.03.24.24304812

**Authors:** Zhengyang Yu, Maohuan Lin, Zhanyu Liang, Bozhen Ren, Ying Yang, Wen Chen, Yonghua Wang, Xiaoling Lin, Yangxin Chen, Kaida Ning, Li C. Xia

## Abstract

Heart failure (HF) is a common cardiovascular syndrome that poses significant morbidity and mortality risks. While genome-wide association studies reporting on HF abound, its genetic etiology remains poorly elucidated, primarily due to its inherent polygenic nature. Furthermore, these genetic insights have not been fully leveraged for the development of effective primary treatment strategies for HF. In this study, we conducted a large-scale integrated multi-trait analysis using European-ancestry GWAS summary statistics of coronary artery disease and HF, involving near 2 million samples to identify novel risk loci associated with HF. 72 loci were newly identified with HF, of which 44 were validated in the replication phase. Transcriptome association analysis revealed 215 HF risk genes, including *EDNRA* and *FURIN*. Pathway enrichment analysis of risk genes revealed their enrichment in pathways closely related to HF, such as response to endogenous stimulus (adjusted P = 8.83×10^-3^), phosphate-containing compound metabolic process (adjusted P = 1.91×10^-2^). Single-cell analysis indicated significant enrichments of these genes in smooth muscle cells, fibroblast of cardiac tissue, and cardiac endothelial cells. Additionally, our analysis of HF risk genes identified 74 potential drugs for further pharmacological evaluation. These findings provide novel insights into the genetic determinants of HF, highlighting new genetic loci as potential interventional targets to HF treatment, with significant implications for public health and clinical practice.

## Introduction

Heart failure (HF) is a common cardiovascular syndrome that causes symptoms, such as shortness of breath, volume overload, and other functional limitations, resulting from structural or functional impairment of ventricular filling or ejection of blood[1]. It poses a significant global health challenge with high prevalence, morbidity, mortality, hospitalization rates, and healthcare costs[2]. Understanding the genetic etiology of HF is crucial, as it can provide insights into the molecular mechanisms underlying the disease, identify novel therapeutic targets, and improve risk stratification through genetic markers. However, the polygenic nature of HF, with numerous small-effect genetic variants, poses significant challenges to fully elucidating its genetic basis.

Current statistical genetic approaches to HF face several limitations. Compared to other cardiovascular diseases, fewer HF-associated loci have been identified, largely due to limited statistical power in genome-wide association studies (GWAS) and smaller sample sizes. Additionally, the genetic architecture of HF is complicated by heterogeneity across subtypes (e.g., HF with preserved ejection fraction vs. reduced ejection fraction) and by the modest availability of diverse population datasets. The presence of linkage disequilibrium further complicates the prioritization of causal variants and genes.

Progress in identifying HF-associated loci has also been slower compared to other cardiovascular diseases, such as hypertension and coronary artery disease (CAD). While hundreds or thousands of loci have been identified for these conditions, HF has only a fraction of validated loci[3–5], partly due to smaller sample sizes and the disease’s clinical heterogeneity. Recent multi-trait genome-wide association studies have shown promise in leveraging shared genetic architectures across related traits, such as stroke and neurodegenerative diseases, to uncover novel loci. For example, multi-Trait Analysis of GWAS (MTAG) has successfully identified novel loci for cardioembolic stroke[6], primary sclerosing cholangitis[7], and Lewy body dementia[8]. Such successes underscore the potential for this methodology to enhance HF research.

To address these challenges, several advanced computational and statistical methods have been proposed. Functional mapping, transcriptome-wide association studies (TWAS), and pathway enrichment analyses integrate multi-omics data to refine variant interpretation. Polygenic risk scoring and fine-mapping approaches enhance the predictive power and resolution of GWAS. However, these methods often require large sample sizes and are sensitive to biases in study design, such as population structure or trait heterogeneity. While these techniques have demonstrated success, their applicability to HF remains constrained by the scarcity of robust datasets and the need for refined analytical frameworks.

MTAG emerges as a particularly powerful tool for analyzing complex diseases like HF. By leveraging genetic correlations among HF and related traits, such as coronary artery disease and hypertension, MTAG enhances statistical power and facilitates the discovery of loci that may not achieve significance in single-trait GWAS. Its ability to distinguish shared from trait-specific genetic effects makes it especially suited for exploring HF’s genetic architecture, where shared pathways with other cardiovascular conditions play a critical role.

HF and CAD, in particular, share a strong genetic correlation, with studies reporting a high genetic overlap (a genetic correlation coefficient of 0.67) [5]. Longitudinal studies have further demonstrated the clinical and genetic interplay between these conditions, such as findings that CAD patients are at elevated risk of developing HF[9]. The growing availability of large GWAS datasets, such as those from the GWAS Catalog[10] and FinnGen[11], offers unprecedented opportunities to study these shared genetic architectures. Additionally, methodological advancements like linkage disequilibrium score regression (LDSC) [12, 13], transcriptome-wide association studies (TWAS)[14–16], and functional annotation tools such as FUMA[17] have further expanded the toolkit for prioritizing causal variants and genes in these complex diseases.

Moreover, drug-repurposing has emerged as an efficient approach to translate approved drugs for new indications based on GWAS-identified risk genes. Currently, although several medications exist for treating HF, most only alleviate symptoms and do not reverse the condition. For HF with preserved ejection fraction, only SGLT2 inhibitors have demonstrated significant reductions in hospitalization and cardiovascular death for patients[18]. Consequently, there is an urgent need to discover new therapeutic drugs for HF, and drug repurposing offers a promising avenue. In recent years, mapping of genome-wide significant GWAS loci to specific genes, thus enabling the reuse of drugs targeting these genes becomes an important repurposing method[19–21]. Another approach involves using TWAS[14, 16] to obtain gene information associated with the phenotype and conducting signature mapping. Specifically, this involves correlating expression level of genes associated with the disease with the pharmacological effects of compounds acting on these genes[22–25].

In this study, we combined MTAG with complementary statistical methods to comprehensively investigate the genetic basis of heart failure (HF). Leveraging the largest European GWAS summary statistics datasets for HF and coronary artery disease (CAD) to date, we identified 99 HF-associated loci, including 72 novel discoveries, 44 of which were validated in a replication phase. Further integration with TWAS uncovered 215 HF risk genes, such as EDNRA and FURIN, newly linked to HF. Pathway enrichment analysis revealed that these genes are involved in critical biological processes, including response to endogenous stimuli, phosphate metabolism, and cell proliferation. Single-cell analysis further localized these genes to smooth muscle cells, cardiac fibroblasts, and endothelial cells, emphasizing their functional relevance. Finally, drug repurposing analysis using MTAG and TWAS findings suggested 74 potential therapeutic candidates for HF, including 21 FDA-approved drugs, offering promising avenues for clinical application and future research.

## Materials and Methods

### Study design, data source and quality control

The complete workflow of this study is illustrated in **Fig. 1**. We obtained the largest GWAS summary statistics of HF (47,309 cases and 930,014 controls), and CAD (181,522 cases and 984,168 controls) from the GWAS Catalog (https://www.ebi.ac.uk/gwas/)[10, 26, 27] for the discovery phase. Additionally, we also obtained GWAS summary statistics for HF (19,350 cases and 288,996 controls) and CAD (33,628 cases and 275,526 controls) from the FinnGen repository https://finngen.gitbook.io/documentation/v/r7/)[11], serving as an independent replication phase. These studies were all conducted on individuals of European ancestry and underwent stringent quality control procedures, as previously described[11, 26, 27].

**Fig. 1.**
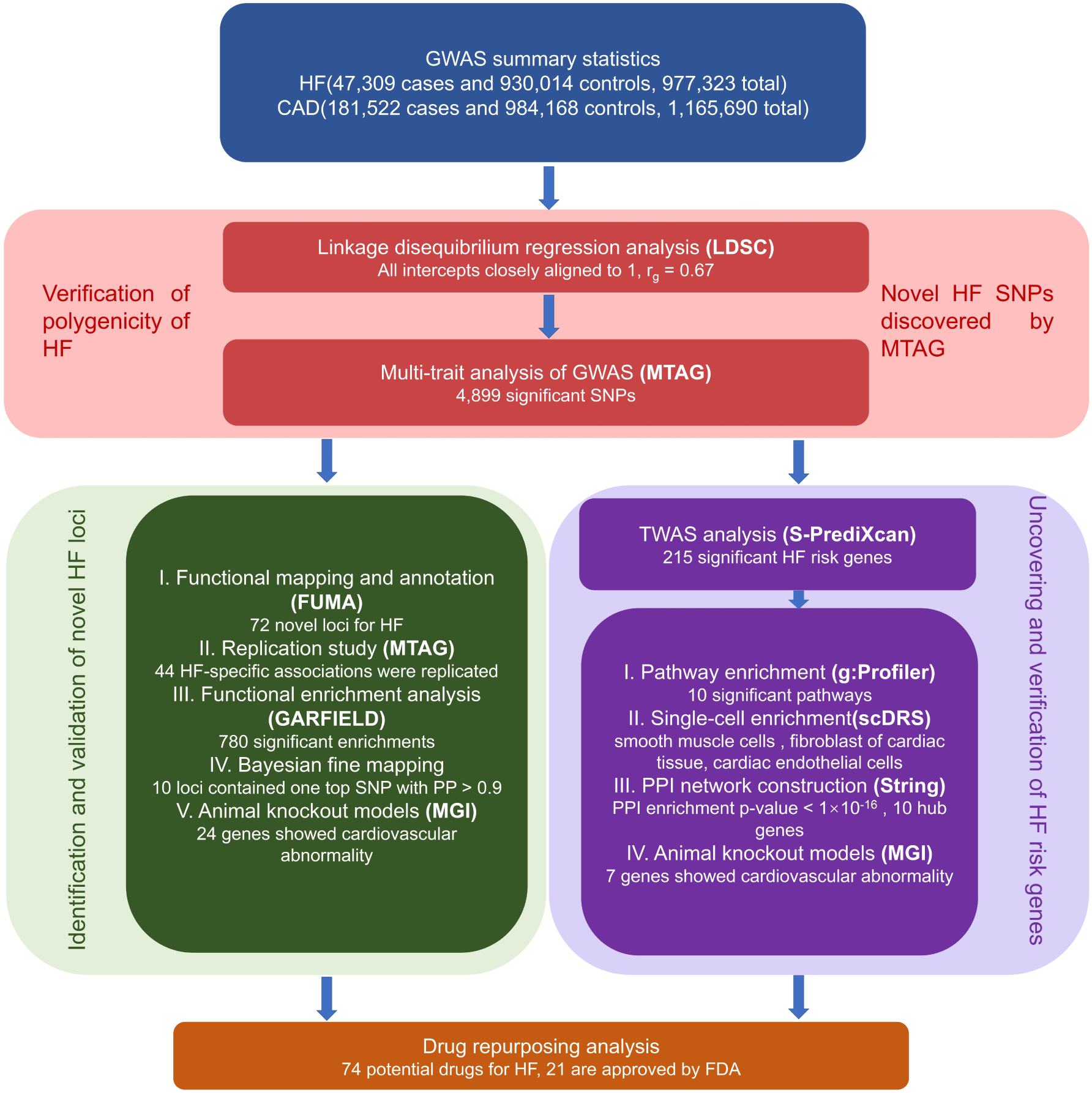
Overall study design. Employing the largest available GWAS summary statistics for heart failure (HF) encompassing 47,309 cases and 930,014 controls, as well as coronary artery disease (CAD) with 181,522 cases and 984,168 controls, we explored the polygenicity and genetic correlation between them using linkage disequilibrium score regression (LDSC), followed by multi-trait analysis of GWAS (MTAG) to identify significant SNPs. Functional annotation at the SNP level was performed to identify novel risk loci paired with MTAG on replication phase to confirm the novel identified risk loci. Additionally, transcriptome-wide association study (TWAS) was conducted to uncover HF risk genes. Subsequently, four gene-based analyses (pathway enrichment, single-cell enrichment, protein-protein interaction network, and animal knockout models) were performed to confirm the role of TWAS risk genes in the development of HF. Finally, drug repurposing was employed to identify potential therapeutic drugs for HF.

In the CAD summary statistics, base pair positions were converted to rs ID, employing the Genome Reference Consortium Human Reference 37 (GRCh37) (<u>Index of /goldenPath/hg19/database(ucsc.edu</u>). Furthermore, we excluded the major histocompatibility complex region (chromosome 6, 26–34 Mb) from our analyses due to its complex structure. SNPs with minor allele frequency less than 0.01 were filtered out, and the analysis was restricted to biallelic SNPs. SNPs with duplicated or missing rs IDs were removed from each GWAS summary dataset for subsequent analyses. A detailed description of all GWAS studies used in this research can be found in **Additional file 1: Table S1**.

### Linkage disequilibrium score regression analysis

To estimate the polygenicity of HF and the genetic correlation between HF and CAD, we used LDSC[12, 13]. We first performed single-trait LDSC analysis on the GWAS summary statistics of HF and CAD to obtain several key parameters, including the mean χ^2^ statistic, genomic inflation factor (λ_GC_), and intercept.

The mean χ^2^ statistic measures the association between genetic variants and a specific trait of interest in the GWAS summary statistics. The traits with a mean χ^2^ statistic < 1.1 were excluded from further analysis. The factor λ_GC_ evaluates the presence of genetic inflation, which refers to an excess of test statistics over the expected null distribution. A value close to 1 indicates that the inflation in test statistics mostly results from polygenic effects. Similarly, the intercept obtained from LDSC analysis (intercept minus one), assesses the contribution of confounding biases to the inflation of test statistics. A value close to 1 suggests that inflation is primarily driven by polygenic effects.

Subsequently, we applied pairwise LDSC analysis to investigate the genetic correlations between HF and CAD. We used the pre-computed linkage disequilibrium scores of European ancestry downloaded from the 1000 Genomes Project Phase 3 (https://alkesgroup.broadinstitute.org/LDSCORE/)[28]. Since that low imputation quality can potentially diminish test statistics, we focused on well-imputed HapMap3 SNPs.

### Multi-trait meta-analysis with MTAG

To boost the power of HF-associated SNP discovery, we used MTAG v.1.0.8[29]. MTAG is a robust method designed to enhance trait-specific power by leveraging genetic correlations among multiple genetically related traits[29]. As input, it takes the GWAS summary statistics of correlated traits and incorporates the LDSC method to estimate the genetic covariance matrix. Subsequently, it generates weights and employs generalized inverse variance weighting to estimate trait-specific effects for a common set of SNPs. MTAG results can be similarly presented as the original GWAS summary statistics for individual traits, providing an easily understood view of genetic architecture.

To replicate newly identified HF-associated loci from MTAG, we also conducted MTAG on HF and CAD from FinnGen repository[11], which are independent from those in discovery phase. In our analyses, the summary statistics derived from single-trait GWAS were denoted as GWAS_HF_ and GWAS_CAD_. Additionally, those acquired from the discovery phase of the MTAG analysis for HF were denoted as MTAG_HF_, while results from the replication phase were denoted as MTAG_HF_R_. The genome-wide significance level for MTAG_HF_ was set at P_MTAG_HF_ < 5 × 10^-8^.

### Functional annotation by FUMA

To investigate the functional implications of HF associated SNPs, we used the Functional Mapping and Annotation (FUMA) platform v1.5.4[17]. Using its default settings, we annotated significant SNPs of MTAG_HF_, using the European ancestry data from the 1000 Genomes Project Phase 3 as a reference. We defined independent significant SNPs as those with P_MTAG_HF_ < 5 × 10^-8^ and independent from each other at r^2^ < 0.6. Lead SNPs are independently significant SNPs that are independent from each other at r^2^ < 0.1. Genomic risk loci were identified by merging linkage disequilibrium blocks of independent significant SNPs in close proximity (< 250 kb). The top lead SNP was recognized as the SNP with the lowest P_MTAG_HF_[17] within a locus.

To gain further insight into the functional annotations of independent significant SNPs and their LD proxies, we employed tools including ANNOVAR categories[30], combined annotation-dependent depletion (CADD) scores[31], and Regulome DB scores[32] with their default parameters. For comparative purposes, we applied the same procedure to genome-wide significant SNPs from GWAS_HF_.

### Bayesian fine-mapping analysis

To prioritize causal SNPs for each locus, we conducted Bayesian fine-mapping analysis to analyze risk loci determined in the FUMA analysis using the *finemap.abf* function of the *coloc v5* R package (https://chr1swallace.github.io/coloc/)[33]. This function leverages bayesian methods to calculate the posterior probability for each SNP being a causal variant within the corresponding risk locus[33]. SNPs were incrementally included into the set based on their descending posterior probabilities, up to the point where cumulative posterior probability reached 0.90, resulting in a 90% credible SNP set for each locus.

### Functional enrichment analysis

To investigate the functional enrichments of significant SNPs of MTAG_HF_, we used GWAS analysis of regulatory or functional information enrichment with LD correction (GARFIELD) [34]. GARFIELD utilizes GWAS summary statistics data, along with regulatory functional annotation data, including genic annotations, histone modifications, transcription factor binding sites, and chromatin segmentation states, across cell types and tissues[34]. Specifically, GARFIELD applies LD pruning based on LD and distance information to select independent SNPs. It then annotates SNPs with regulatory information if they, or highly correlated SNPs with them, overlap with such regulatory features. Finally, it computes odds ratios (OR) and enrichment p-values using logistic regression. We determined the enrichment to be significant at Bonferroni-corrected p < 4.98×10^-5^ (0.05/1005), where 1,005 represents the number of feature annotations. We also applied GARFIELD to GWAS_HF_ for comparison.

### Transcriptome-wide association analysis

To fully harness the information from SNPs for prioritizing risk genes associated with HF, we used the S-PrediXcan method[16] combined with the Joint Transcriptome Imputation (JTI) model[35] to perform TWAS analysis on MTAG_HF_. S-PrediXcan integrates summary-level GWAS data with gene expression prediction models, providing an estimate of the association between gene expression and HF[36]. To strengthen the accuracy and robustness of S-PrediXcan analysis, we applied the JTI model. This model combines information from multiple tissue-specific gene expression prediction models, enabling more effective integration of gene expression data across tissues[35]. We used JTI models derived from the Genotype-Tissue Expression (GTEx) project version 8 transcriptome data[37], for eight different tissues associated with heart. These tissues include subcutaneous adipose tissue, visceral omental adipose tissue, atrial appendage, left ventricle, kidney cortex, liver, lung, and whole blood. We applied a Bonferroni correction for multiple testing in each tissue. We also conducted S-PrediXcan on GWAS_HF_ as a comparison.

### Functional characterization and contextual analysis for risk genes

To gain insight into the biological processes associated with risk genes identified from TWAS, we conducted pathway enrichment analysis using the web-based tool g:Profiler[38]. Pathways were considered significant if they reached a Bonferroni-corrected significance threshold (adjusted P < 0.05). For comparative purposes, we conducted separate enrichment analyses on the risk genes identified by TWAS on GWAS_HF_.

We employed the scDRS method[39] to explore the association between the expression levels of HF risk genes and cardiac-relevant cells. scDRS integrates single-cell RNA sequencing data with gene-disease association information derived from GWAS[39]. It links individual cells to disease status by assessing the excess expression of HF risk genes within each cell. We also applied the scDRS method to risk genes identified by TWAS on GWAS_HF_ as a comparative analysis.

Furthermore, we investigated the functional enrichment of PPI networks among risk genes identified from TWAS via the STRING v12.0 database[40]. We then used the cytoHubba plugin in cytoscape to determine the top ten genes as hub genes[41].

### Querying the MGI database

To confirm the roles of novel genes identified by MTAG and hub genes in our study within cardiovascular diseases, we queried the Mouse Genome Informatics (MGI, http://www.informatics.jax.org/) resource[42] for information on knockout models associated with them. MGI is a pivotal bioinformatics resource, providing comprehensive datasets pertaining to the mouse genome, and it is comprised of gene functionalities, expression patterns, mutation profiles, and data related to disease models. We focused on genes for which cardiovascular abnormalities became apparent after knockout experiments[42].

### Drug repurposing analysis

In order to translate our genetic findings into therapeutic strategies for treating HF, we conducted drug repurposing, with the main steps outlined as follows: We first selected all TWAS risk genes associated with HF and genes identified by MTAG. Subsequently, we utilized the GEN2FUNC function within FUMA to map these genes onto the DrugBank database to generate a preliminary list of candidate drugs[43]. We then obtained a list of 26 FDA-approved HF drugs from Drug Bank[44] (**Additional file 1: Table S2**). For each candidate drug, we calculated its similarity value to all FDA-approved HF drugs, and subsequently averaged these values to obtain an overall similarity score. Here, similarity between two drugs is calculated using DICE score based their molecular fingerprints using drug structure data from DrugBank v5.1.10[44]. Finally, we identified drugs with an average similarity value greater than 0.2 as potential HF drugs. As a comparison, we also performed drug repurposing using 27 reported MTAG genes and the risk genes identified through TWAS results on GWAS_HF_.

## Results

### LDSC analysis revealed the polygenicity of HF and CAD, along with a strong genetic correlation between them

Single-trait LDSC analysis suggested that the inflation observed in the test statistics is likely a consequence of polygenicity, rather than population stratification (**Additional file 1: Table S3)**. The mean χ^2^ statistics were both greater than 1.1. LDSC intercepts consistently remained close to 1. Pairwise LDSC analysis further revealed a strong positive genetic correlation between HF and CAD (r_g_ = 0.67, se = 0.03; P = 8.03×10^-112^). These findings supported our rationale for conducting a multi-trait analysis of HF, CAD.

### MTAG analysis identified new HF-associated risk loci with evidence of replication

We conducted MTAG on GWAS_HF_ and GWAS_CAD_, and identified 4,899 SNPs significantly associated with HF (P_MTAG_HF_ < 5×10^-8^), among which 4,749 SNPs did not reach genome-wide significance in the original GWAS_HF_ analysis (**Fig. 2a, b**).For instance, rs6841581, located within the exon of the *EDNRA* gene, demonstrated an increased significance in MTAG analysis (P_MTAG_HF_ = 4.75×10^-18^) compared to GWAS_HF_ analysis (P_GWAS_ = 1.41×10^-2^) to. Similarly, rs3918226, located within the intron of the *NOS3* gene, exhibited an increase in significance from GWAS_HF_ analysis (P_GWAS_ = 5.98×10^-5^) to MTAG_HF_ analysis (P_MTAG_HF =_ 1.09×10^-22^). From the original GWAS results to multi-trait results, the mean χ^2^ statistics increased from 1.158 to 1.340. These results suggested MTAG analysis could be more powerful than single-trait analysis by leveraging the genetical correlation among multiple traits. And the maxFDR for MTAG_HF_ is 0.03, suggesting no overall inflation due to violation of the homogeneous assumption.

**Fig. 2.**
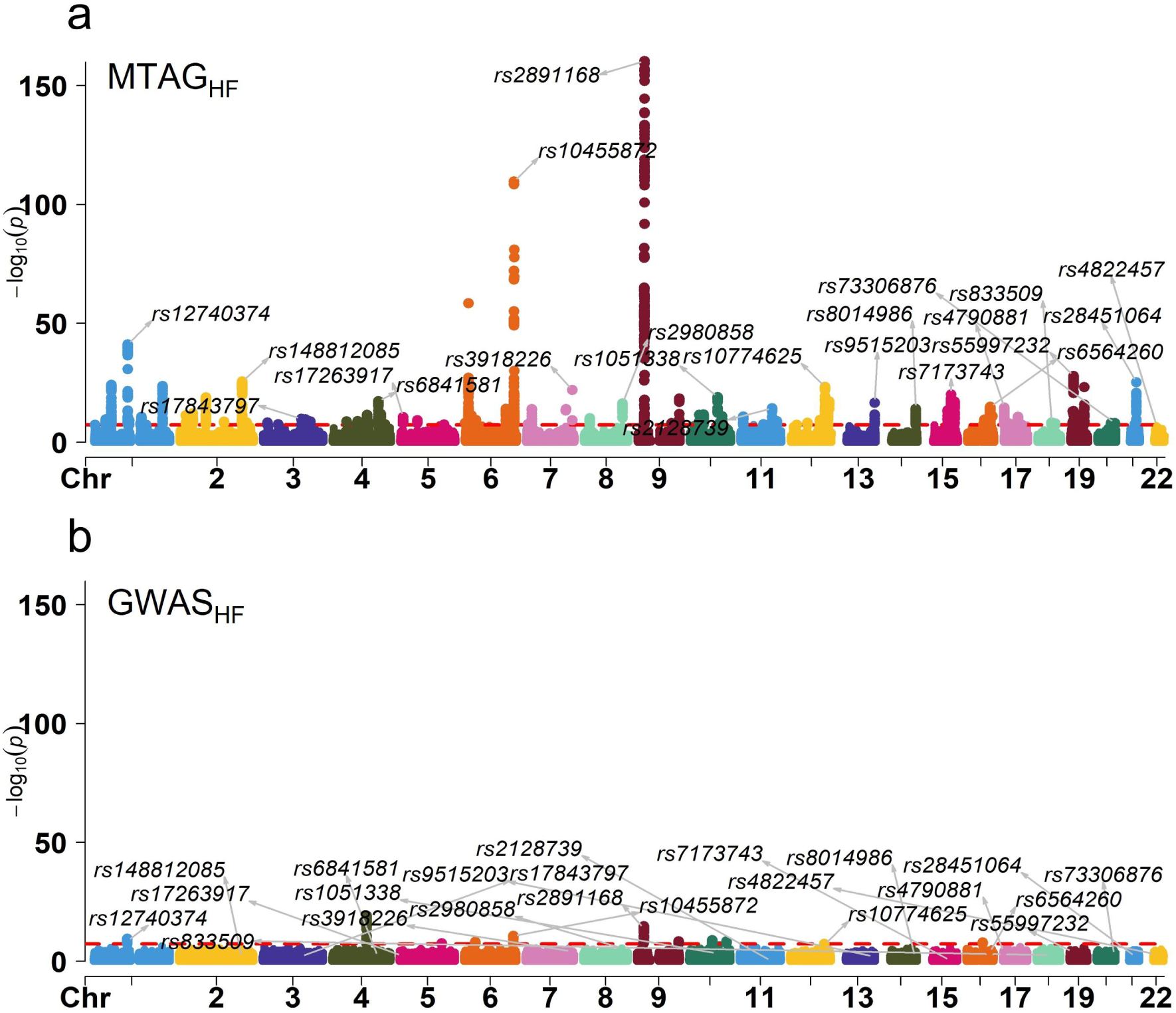
MTAG analysis boosted SNP discovery power. Manhattan plots for MTAGHF (a) and GWASHF (b). Horizontal red dashed lines indicate the genome-wide significance threshold (P < 5 × 10-8). SNPs are labeled in place on chromosome coordinates.

Through FUMA annotation of the 4,899 significant SNPs, we pinpointed 99 risk loci **(Additional file 2: Fig. S1)**. Among these loci, 27 had been previously reported, while 72 were newly discovered. To verify these newly identified risk loci, we performed MTAG analysis on GWAS summary statistics data obtained from the FinnGen repository[11]. Notably, these summary statistics were independent from the GWAS conducted in the discovery phase.

We replicated 44 HF-specific associations at the nominal significance level of 0.05 **(Additional file 1: Table S4)**.Notable examples included the previously mentioned rs6841581 in *EDNRA* (P_MTAG_HF_R_ = 8.01×10^-9^) and rs3918226 in *NOS3* (P_MTAG_HF_R_ = 4.22×10^-5^). Additionally, other associations such as rs7173743 in *MORF4L1* (P_MTAG_HF_R_ = 2.36×10^-4^), rs12509595 in *FGF5* (P_MTAG_HF_R_ = 3.34×10^-3^), rs2306556 in *GUCY1A3* (P_MTAG_HF_R_ = 5.60×10^-3^), and rs1250258 in *FN1* (P_MTAG_HF_R_ = 4.08×10^-2^) were also successfully replicated.

### Functional annotation for significant SNPs of MTAG_HF_

We conducted FUMA to investigate the functional characteristics of significant SNPs (**Additional file 1: Table S5**). For candidate SNPs located within risk loci, most (97.42%) were located in non-coding regions, including UTR3, UTR5, downstream, intergenic region, intron, ncRNA intron and upstream. Only a small proportion of SNPs were exon with 144 (1.41%) in coding RNA and 116 (1.17%) in non-coding RNA (**Fig. 3a**). Among exonic SNPs in coding RNA, rs10965215 was the most statistically significant (P_MTAG_HF_ = 2.87 × 10^-58^) and was mapped to the *CDKN2B-AS1* gene. This was followed by rs3798220 (P_MTAG_HF_ = 6.74 × 10^-51^), which was mapped to the *LPA gene*. The most significant exonic SNP in non-coding RNA was rs564398 (P_MTAG_HF_ = 1.95 × 10^-54^), mapped to the *CDKN2B-AS1* gene.

**Fig. 3.**
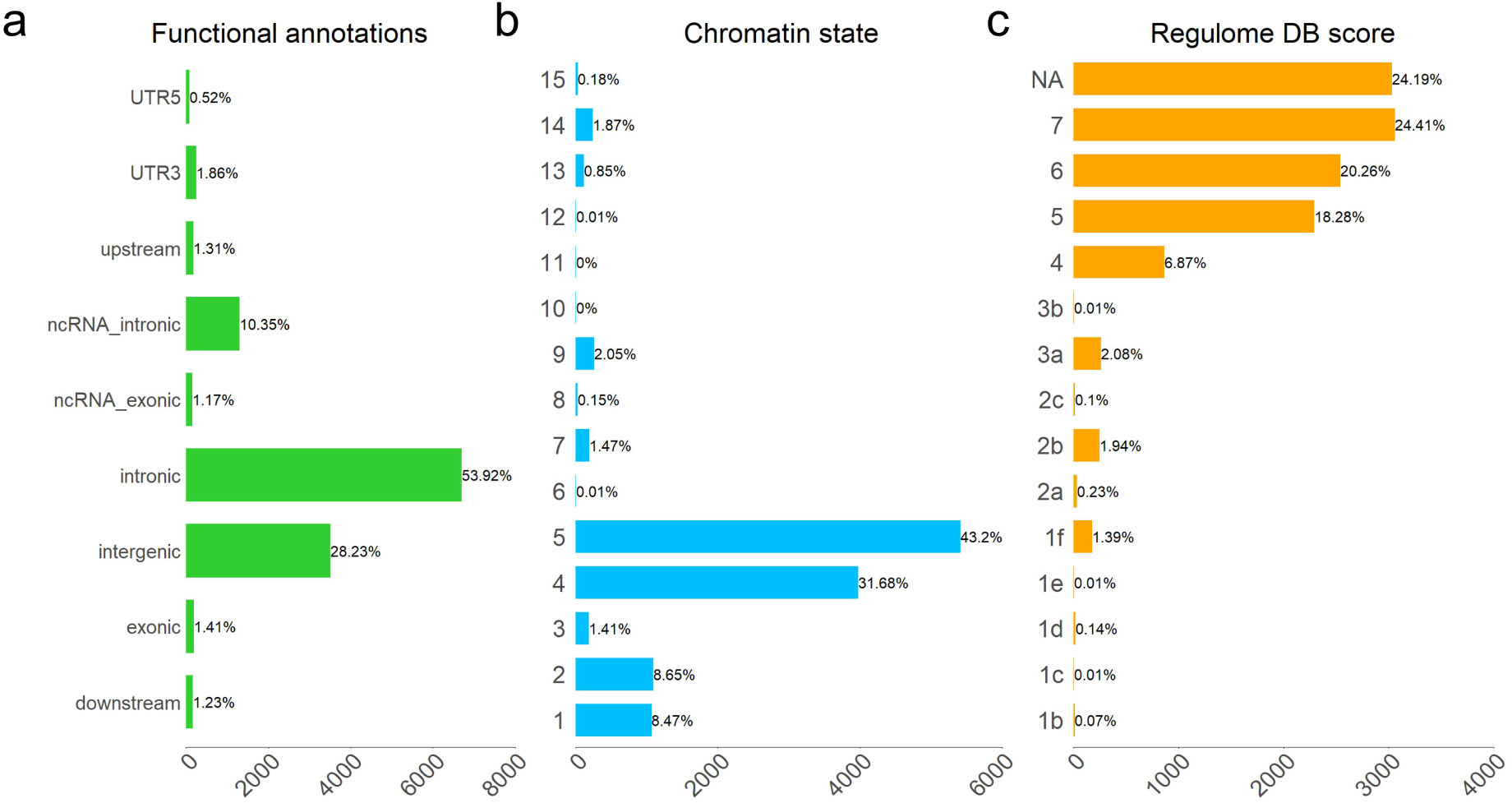
Functional annotations of HF-associated SNPs. (a) Functional annotation of candidate SNPs. (b) Distribution of candidate SNPs across 15 categories of minimum chromatin state. Chromatin states with a value ≤ 7 are considered as open chromatin regions. (c) RegulomeDB scores of candidate SNPs. RegulomeDB scores from 1b to 2c are considered likely to affect transcription factor binding.

Furthermore, 94.89% of candidate SNPs were located in open chromatin regions (minimum chromatin state ≤ 7) (**Fig. 3b**), 3.89% of candidate SNPs were likely to affect the binding of transcription factors (RegulomeDB scores from 1b to 2c) (**Fig. 3c**), and 3.62% of candidate SNPs were deemed potentially deleterious (Combined Annotation Dependent Depletion score > 12.37). Notably, the SNP with the highest CADD score was rs328 (CADD score 50) located in the 8p21.3 locus. It is an exonic SNP of gene *LPL*. All afore mentioned genes (*LPA, CDKN2B-AS1,* and *LPL*) have been associated with HF[45] and CAD[46, 47] in previous studies, now with higher-level of evidence offered in this study.

### Defining SNP credible sets within risk loci

To facilitate the shortlisting of causal SNPs for each of the 99 risk loci, we identified 1,982 SNPs within the 90% credible set using Bayesian fine mapping test (see **Methods**, **Additional file 1: Table S6**). The credible set of 10 HF loci contained only the top SNP with posterior probability greater than 0.90. The credible set of 85 HF loci contained multiple SNPs. The credible set of the remaining four risk loci (1p36.32, 4q33, 5q31.3, 21q22.12) contained no SNPs.

Among the 10 loci with exactly one top SNPs, two were particularly noticeable. One was the top SNP rs3918226 (P_MTAG_HF_ = 1.09×10^-22^) in locus 7q36.1. This SNP was associated with systolic blood pressure[48] and myocardial infarction[49], and it was an intron of the *NOS3* gene, with a CADD score of 12.89. The protein encoded by the NOS3 gene synthesizes endothelial nitric oxide, modulates vascular function, and maintains cardiovascular health. Research has also revealed that mice with a knockout of the *NOS3* gene exhibit cardiovascular abnormalities, including increased heart weight and abnormal cardiac muscle relaxation[50].

The other top SNP was rs7412 (P_MTAG_HF_ = 6.95×10^-24^) in locus 19q13.32. It was within an exon of the *APOE* gene and possessed a high CADD score of 25.1. Additionally, it has been associated with low-density lipoprotein cholesterol measurement[51], triglycerides[52], and pulse pressure[48]. The *APOE* gene encodes a protein known as apolipoprotein E, which plays a crucial role in lipid metabolism, including cholesterol transport and lipoprotein clearance, thereby influencing cardiovascular health. Study has indicated that the knockout of *APOE* in mice increases the risk of atherosclerosis in arteries[53].

### Functional enrichment of significant MTAG_HF_ SNPs

We observed 780 significant enrichments of significant MTAG_HF_ SNPs in regulatory and functional categories **(Additional file 1: Table S7)**, As a comparison, among the significant SNPs of GWAS_HF_ results, only 3 reached the significance threshold **(Additional file 1: Table S8)**. Regarding genic regions (**Fig. 4a**), the most significant enrichment of SNPs was found in exonic regions (OR = 3.76, P = 2.34×10^-10^). In terms of tissue specific DNase I hypersensitive sites (**Fig. 4b**), SNPs were significantly enriched in tissues, such as fetal heart, blood vessel, fetal large intestine, gingival, embryonic stem cell, and fetal small intestine, with fetal heart being the most significant (OR = 4.94, P = 6.14×10^-18^). Concerning tissue-specific chromatin states (**Fig. 4c**), SNPs were significantly enriched in the transcribed region, with the most significant enrichment found in the actively transcribed region of liver tissue (OR = 2.67, P = 1.00×10^-8^). In terms of tissue specific histone-modified regions, SNPs showed a significant enrichment across tissues, such as blood, liver, and cervix (**Fig. 4d**), with the H3K4me1 region of blood tissues (OR = 3.22, P = 2.45×10^-11^) being the most significant.

**Fig. 4.**
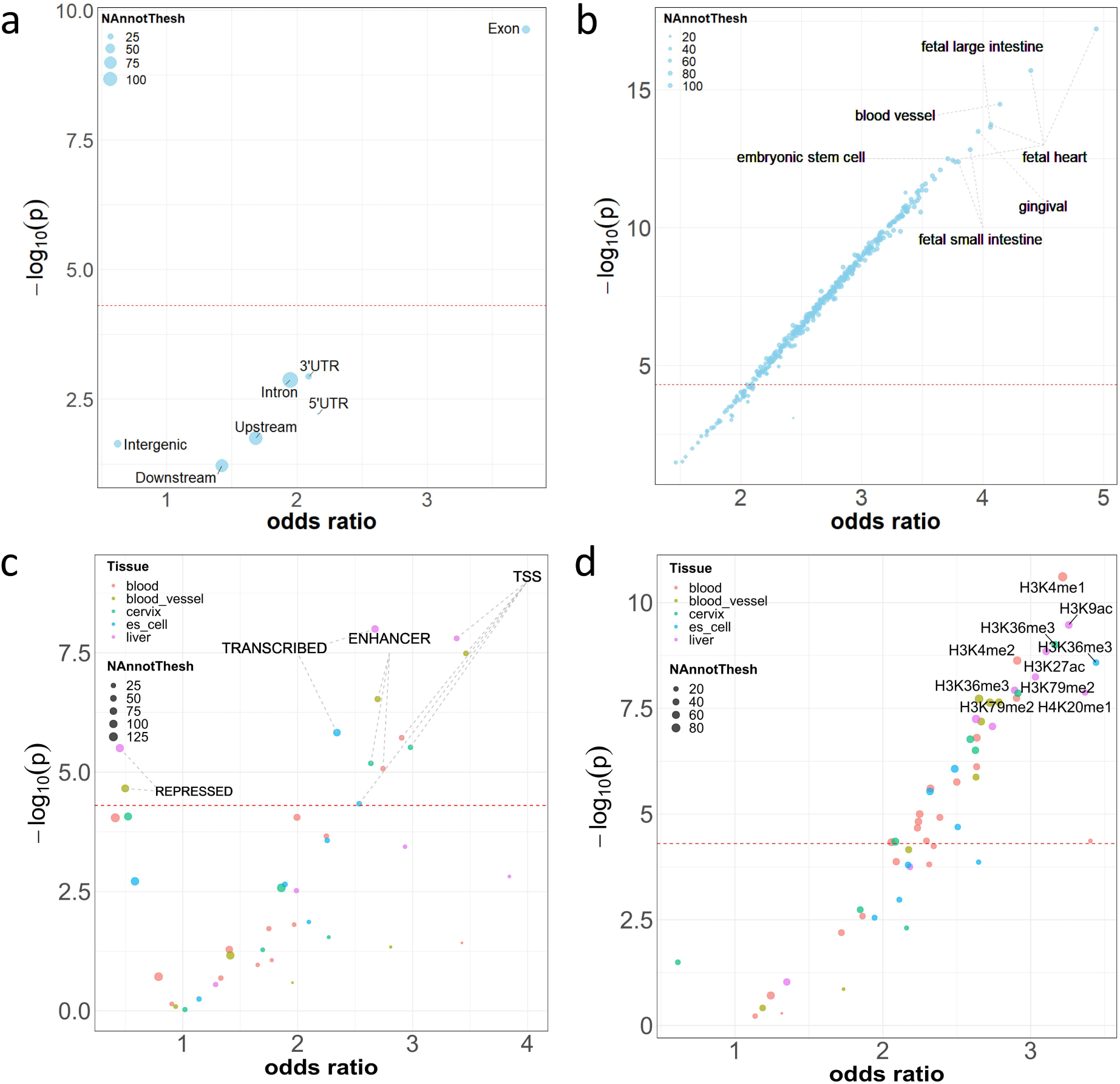
Functional enrichment of significant MTAG_HF_ SNPs. A total of 4,899 SNPs were analyzed with GARFIELD. Enrichment by **(a)** genic regions, **(b)** tissue-specific DNase I hypersensitive sites (DHS) regions **(c)** tissue-specific chromatin states, and **(d)** tissue-specific histone-modified regions. In all plots, enrichment odds ratios were plotted against −log_10_(P value). The dotted horizontal lines mark the statistical significance threshold of P = 0.05/1005.Size of markers is proportional to the number of independent significant SNPs in a specific annotation category, while in subplots **(c)** and **(d)**, markers are also color-coded by tissue categories.

### Uncovering HF risk genes through TWAS

To uncover HF risk genes for subsequent in-depth functional analysis, we conducted a comprehensive transcriptome-wide association analysis (TWAS) on MTAG_HF_ (see **Methods**). After Bonferroni correction to adjust for multiple testing within each tissue, we identified 215 genes that displayed significant associations with HF in at least one tissue type (**Additional file 1: Table S9**).

Among these genes, two were noteworthy. The first *was EDNRA* (P_TWAS_ = 1.72×10^-17^ in adipose visceral omentum tissue), which was previously linked to ischemic stroke[54] and pulse pressure[55], but was newly associated with HF in this study. Importantly, it was absent from the TWAS results based solely on GWAS_HF_ **(Additional file 1: Table S10)**. *EDNRA* encodes the Endothelin Receptor Type A, which is a G protein-coupled receptor predominantly expressed in endothelial cells and smooth muscle cells. It plays a pivotal role in mediating the signaling of Endothelin-1 hormone. Research suggested that the deficiency in this gene could result in cardiovascular anomalies in mice, such as elevated blood pressure, abnormal cardiovascular system morphology, and the dilation of the right ventricle[56].

The second gene of significance is *FURIN*. This study was the first to report the association between *FURIN* and HF. *FURIN* (P_TWAS_ = 3.09×10^-15^ in liver tissue) is a gene encoding a protein known as furin, which belongs to the family of serine proteases. This protease primarily functions within the cell’s Golgi apparatus and vesicles, playing a pivotal role in the post-translational modification and processing of proteins. Research has indicated that the absence of furin leads to severe cardiovascular anomalies, including anomalous cardinal vein morphology, the absence of vitelline blood vessels, and abnormal heart development in developing mouse embryos[57].

### Contextual analysis of TWAS risk genes

We obtained 10 significantly enriched pathways of TWAS risk genes (**Fig. 5a**, **Additional file 1: Table S11**). These pathways primarily involve metabolic processes of important compounds, and cell proliferation. such as response to endogenous stimulus (adjusted P = 8.83×10^-3^), phosphate-containing compound metabolic process (adjusted P = 1.91×10^-2^), and cell population proliferation (adjusted P = 2.41×10^-2^). In comparison, risk genes identified by GWAS_HF_ showed no enrichment in any pathway.

**Fig. 5.**
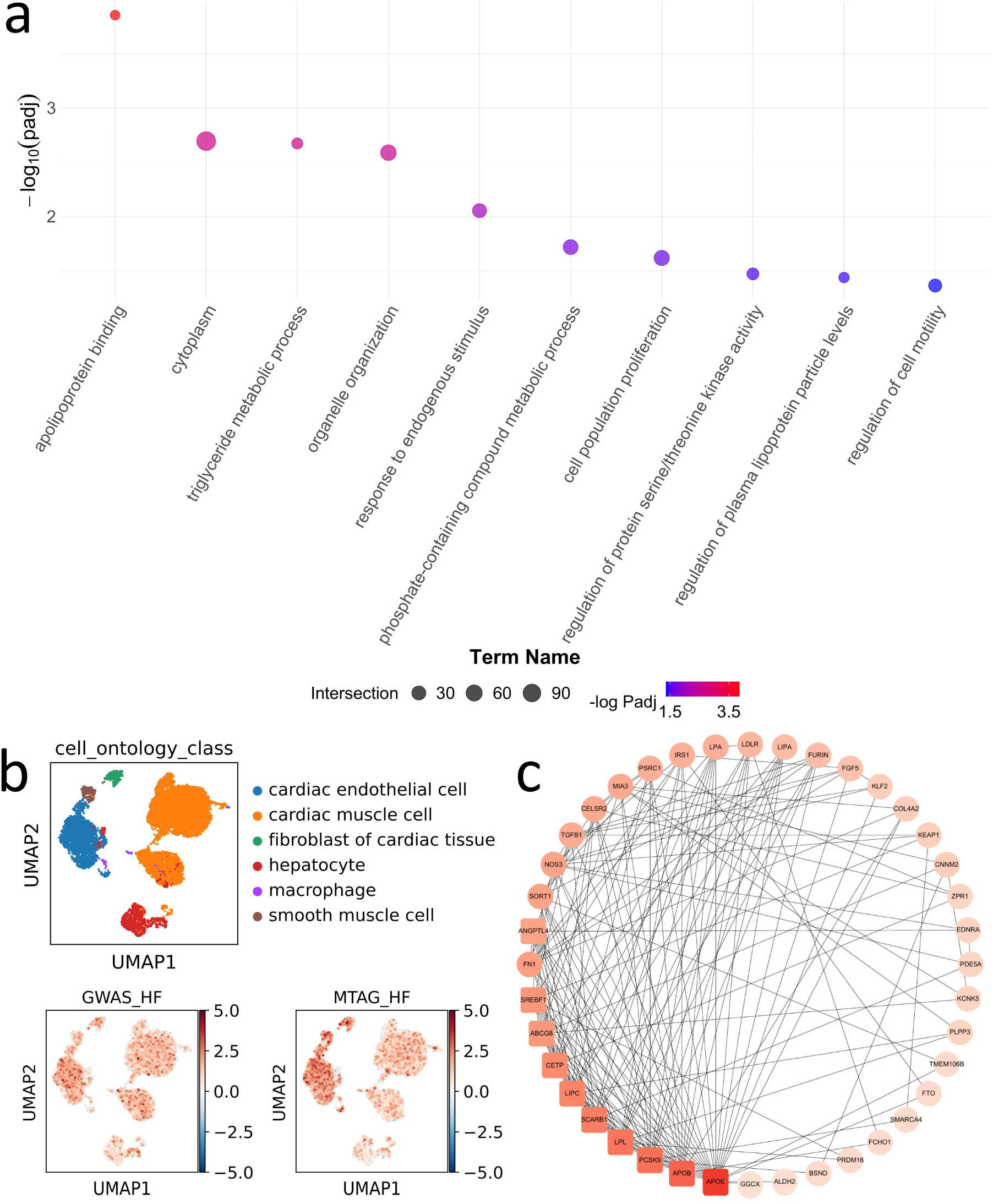
Contextual analysis of TWAS risk genes. **(a)** Bubble plot of 10 enriched pathways. The x-axis represents pathway names, and the y-axis represents-log_10_(padj). Size of the bubbles indicates the number of genes enriched in the corresponding pathway, while the color of the bubbles changes according to the-log_10_(padj). **(b)** Enrichment of risk genes at the cellular level. Node color represents the risk score on the cells with red indicating high risk and blue indicating low risk. **(c)** Protein-protein interaction (PPI) sub-network of 10 hub genes. Gene nodes are arranged from left to right based on their degrees, with round rectangle representing the hub genes.

Using single-cell enrichment analysis, we found that HF risk genes were enriched in smooth muscle cells, fibroblast of cardiac tissue, cardiac endothelial cells (**Fig. 5b, Additional file 1: Table S12**). Risk genes identified by GWAS_HF_ showed no enrichment in any cardiac cells **(Additional file 1: Table S13)**.

We also constructed a protein-protein interaction (PPI) network on the risk genes, resulting in a network with 204 nodes and 402 edges (**Additional file 1: Fig. 2**). The PPI network demonstrated significant enrichment (P < 1×10^-16^), indicating that the proteins in this network are biologically interconnected. Among all genes, ten hub genes were identified: *APOE, APOB, PCSK9, LPL, SCARB1, LIPC, CETP, ABCG8, SREBF1, ANGPTL4* (**Fig. 5c**).

### Mouse knock-out models for novel MTAG genes and hub genes

We conducted queries in silico knock out mouse models using the Mouse Genomics resource to find evidence of target gene modifications that could produce phenotypes associated with HF (**Additional file 1: Table S14**). A total of 29 gene knock-out models yielded evidence associated with cardiovascular abnormalities. Specifically, knock-out models of *PRDM16, ABCG8, FN1, FGD5, EDNRA, GUCY1A3, NOS3, PLCE1, CNNM2, MORF4L1, FES* showed evidence for congestive heart failure, myocardial abnormalities, cardiac hypertrophy, and abnormal myocardial fiber morphology, all of which were related to the development of HF.

### Drug repurposing analysis

We identified 74 potential therapeutic drugs based on MTAG genes and HF risk genes identified by TWAS (see **Methods**, **Additional file 1: Table S15**), 21 of which were approved by the U.S. FDA (**Table 1**). Notably, five (DB06403, DB00945, DB00559, DB08932, and DB06268) have undergone clinical trials for the treatment of HF[58–62]. Another three (DB09237, DB05676, and DB12548) have been subjected to clinical trials for diseases related to HF, such as hypertension[63, 64] and vascular inflammation[65]. The remaining 53 candidate drugs are investigational compounds that have not been approved by the U.S. FDA for treatment of any disease. In contrast, using 27 reported GWAS genes and the risk genes identified through TWAS results on GWAS_HF_, we identified only seven drug compounds. However, their overall similarity score to FDA-approved HF drugs were all below 0.2 and did not pass the threshold for repurposing (**Additional file 1: Table S16**).

**Table 1.** Twenty-one FDA-approved drugs were identified as candidate drugs for HF.

| DrugBank ID | Generic Name | Target | Similarity score |
| --- | --- | --- | --- |
| DB09237 | Levamlodipine | *NOS3* | 0.346 |
| DB01125 | Anisindione | *GGCX* | 0.311 |
| DB06237 | Avanafil | *PDE5A* | 0.305 |
| DB05676 | Apremilast | *NOS3* | 0.291 |
| DB06403 | Ambrisentan | *EDNRA* | 0.291 |
| DB12548 | Sparsentan | *EDNRA* | 0.285 |
| DB12010 | Fostamatinib | *FES* | 0.272 |
| DB04951 | Pirfenidone | *FURIN* | 0.269 |
| DB00945 | Aspirin | *EDNRA* | 0.261 |
| DB00820 | Tadalafil | *PDE5A* | 0.254 |
| DB00559 | Bosentan | *EDNRA* | 0.252 |
| DB08932 | Macitentan | *EDNRA* | 0.248 |
| DB06589 | Pazopanib | *SH2B3* | 0.248 |
| DB00170 | Menadione | *GGCX* | 0.246 |
| DB09332 | Kappadione | *GGCX* | 0.242 |
| DB01110 | Miconazole | *NOS3* | 0.238 |
| DB06268 | Sitaxentan | *EDNRA* | 0.231 |
| DB06267 | Udenafil | *PDE5A* | 0.229 |
| DB00862 | Vardenafil | *PDE5A* | 0.217 |
| DB00203 | Sildenafil | *PDE5A* | 0.215 |
| DB01022 | Vitamin K1 | *GGCX* | 0.200 |

## Discussion

In this study, we conducted a multi-trait analysis by combining HF and CAD, leading to the identification of 72 novel HF risk loci and 215 HF risk genes through TWAS. Based on these genetic discoveries, we conducted drug repurposing and identified 74 potential therapeutic drugs for HF, among which 21 were U.S. FDA-approved.

Using summary statistics data from the largest HF and CAD GWAS, we proved a highly positive genetic correlation between HF and CAD, which was corroborated by previously findings[4]. This connection led us hypothesize that these cardiovascular diseases with high genetic correlation could be analyzed with the multi-trait approach to expand novel genetic discoveries by integrating large-scale data[6, 66, 67].

Our multi-trait analysis significantly improved the identification of HF risk loci. Specifically, we identified 72 novel loci associated with HF. One noteworthy risk locus is the 2p21, with the top lead SNP rs4245791 reaching significance in GWAS_CAD_[26] and demonstrating a consistent effect direction with MTAG_HF_. The locus mapped to the gene *ABCG8*, also known as ATP binding cassette subfamily G member 8. *ABCG8* encodes a protein crucially involved in cholesterol transport and homeostasis. Research has revealed that its deficiency in mice lead to abnormalities in heart left ventricle morphology, along with an increase in cardiac muscle contractility[68]. Another noteworthy risk locus is 15q25.1, with the top lead SNP being rs7173743. The SNP was an intronic of gene *MORF4L1*. *MORF4L1*, also known as Mortality Factor 4 Like 1, is a gene responsible for encoding a protein engaged in cellular processes, such as chromatin remodeling and DNA repair. Research has additionally demonstrated that its deficiency in mice resulted in cardiovascular abnormalities, including myocardial fiber disarray and cardiac hypertrophy[69].

Since the replication analysis is recommended to evaluate the credibility of each SNP association when applying MTAG to low-powered GWAS or to GWAS characterized by significant heterogeneity in statistical power. We conducted MTAG analysis on the FinnGen cohort serving as the replication phase. Our findings revealed that out of the 99 loci, 68 exhibited replicated HF-specific associations. Additionally, among the 72 novel loci identified, 44 were successfully replicated. Notably, several genes mapped with replicated loci showed direct relevance to cardiovascular abnormalities, such as myocardial abnormalities, cardiac hypertrophy, and abnormal myocardial fiber morphology, in mouse knockout models. These genes include *EDNRA*, *NOS3*, *MORF4L1*, *FGF5*, *GUCY1A3*, and *FN1*.

We further conducted a TWAS analysis and identified 215 HF risk genes and validated the roles of these genes in the progression of HF. Pathway enrichment analysis identified several key pathways closely associated with HF, including response to endogenous stimulus, phosphate-containing compound metabolic process, and cell population proliferation. Response to endogenous stimulus refers to any process that induces a modification in the state or function of a cell or organism, encompassing alterations in movement, secretion, enzyme production, gene expression, and other physiological activities. Further, a close association of endogenous hemodynamic[70] and endogenous nitric oxide[71] with HF has been reported. The phosphate-containing compound metabolic process pathway encompasses chemical reactions related to the phosphate group, which is the anion or salt derived from any phosphoric acid. This includes processes such as the synthesis of adenosine triphosphate. Notably, a reduction in the energy reserve available for adenosine triphosphate synthesis can render the heart more susceptible to systolic and diastolic failure[72]. The cell population proliferation pathway refers to the multiplication or reproduction of cells, leading to the expansion of a cell population. The proliferation of cardiac myocytes is intricately linked to the development of HF[73–75], and contemporary researches have proposed various approaches aimed at inducing cardiac myocyte proliferation as a therapeutic strategy for HF[76–78].

In the context of single-cell enrichment, it is noteworthy that risk genes demonstrated enrichment in cell types, such as smooth muscle cells, fibroblasts of cardiac tissue, and cardiac endothelial cells. Additionally, an earlier study has demonstrated that the deletion of the mineralocorticoid receptor specifically from smooth muscle cells can mitigate HF induced by transverse aortic constriction[79]. Furthermore, contemporary research indicated that endothelial cells play a crucial role in both coronary microvascular dysfunction and cardiac remodeling, ultimately contributing to the development of HF[80]. Moreover, investigations have highlighted the involvement of cardiac fibroblasts in myocardial stiffening through collagen production, as well as their active participation in modulating cardiac inflammation by synthesizing chemoattractive substances[81].

Drug repurposing facilitated by GWAS methods has gained popularity in recent years[82]. Our study extensively leveraged the genetic discoveries including those loci identified by MTAG and the risk genes identified through TWAS to explore potential drugs for primary prevention of HF. We identified 74 potential therapeutic drugs for HF treatment. Notably, five of these approved drugs (DB06403, DB00945, DB00559, DB08932, and DB06268) have undergone clinical trials for the treatment of HF[58–62], thus boosting the credibility of our findings. Moreover, the target genes of these potential medications exhibited substantial evidence of associations with cardiovascular system abnormalities in knock out models. Notably, *ALDH2*, *NOS3*, *MIF*, *FURIN,* and *PDGFD*, demonstrated direct associations with such pathological features as congestive heart failure, myocardial abnormalities, cardiac hypertrophy, and abnormal myocardial fiber morphology[50, 57, 83–85]. Furthermore, *ALDH2*, *EDNRA*, and *FURIN*, which exhibited expression levels positively correlated with HF risk, may represent more favorable targets for the development of novel therapies, since therapeutics aimed at downregulating a gene are typically easier to develop than those aimed at upregulation. It is noteworthy that using solely previously reported loci information and the original GWAS_HF_ data, we failed to identify any drugs.

Despite these novel findings, our study had certain limitation. Our data were derived from European populations, thus potentially limiting the generalizability of our study to other ethnicities. However, recent advancements have introduced methods for transferring genetic findings from European populations to others, increasing the applicability of our research[86, 87].

## Conclusions

In conclusion, our study identified 72 novel HF risk loci. Subsequently, we employed MTAG on replication phase, Bayesian fine mapping, GARFIELD, and MGI queries to establish the biological and statistical credibility of these loci. We also identified 215 significant HF risk genes through TWAS analysis. Pathway enrichment, single-cell enrichment, PPI network, and MGI queries confirmed the roles of these genes in the progression of HF. Based on these novel HF genes, we identified 74 drugs suitable for repurposing for HF. These findings have translational value in future efforts aimed toward the discovery of HF prevention and treatment regimens.

## List of abbreviations

HF: Heart failure
GWAS: Genome-wide association study
CAD: Coronary artery disease
LDSC: Linkage disequilibrium score regression
MTAG: Multi-trait analysis of GWAS
SNP: Single nucleotide polymorphism
TWAS: Transcriptome-wide association study
GTEx: Genotype-Tissue Expression
GRCh37: Genome Reference Consortium Human Reference 37
FUMA: Functional mapping and annotation of genetic associations
CADD: Combined annotation-dependent depletion
GARFIELD: GWAS analysis of regulatory or functional information enrichment with linkage disequilibrium correction
JTI: Joint-tissue imputation
MGI: Mouse Genome Informatics
OR: Odds ratio
PPI: Protein-protein interaction.

## Declarations

### Ethics approval and consent to participate

Not applicable.

### Consent for publication

Not applicable.

### Availability of data and materials

The datasets analysed during the current study are publicly available. GWAS summary statistics data for discovery phase are available from GWAS Catalog under study accessions GCST009541[88] and GCST90132314[89] at https://www.ebi.ac.uk/gwas/. GWAS summary statistics data for replication phase are available at https://storage.googleapis.com/finngen-public-data-r7/summary_stats/finngen_R7_I9_HEARTFAIL_ALLCAUSE.gz and https://storage.googleapis.com/finngen-public-data-r7/summary_stats/finngen_R7_I9_CHD.gz[11]. Data used in LDSC analysis can be obtained at https://alkesgroup.broadinstitute.org/LDSCORE/[28]. JTI models of gene expression in 8 heart tissues are available from Zenodo at http://doi.org/10.5281/zenodo.3842289[90]. Single-cell data used in scDRS analysis are available from Tabula Sapiens Single-Cell Dataset at https://doi.org/10.6084/m9.figshare.14267219.v5[91]. All drug information is available from Drugbank v.5 at https://go.drugbank.com/[92]. The information on all mouse knockout models can be obtained from Mouse Genome Informatics resources at https://www.informatics.jax.org/[93].

### Authors’ contributions

Z.Y., Y.C., K.N., and L.C.X. conceived and designed the study. Z.Y. and Z.L. collected the data. Z.Y. and Z.L. performed the data analysis. Z.Y., Y.Y., B.R. and M.L. conducted the interpretation of analysis results. Z.Y., L.C.X, and K.N. wrote the manuscript. Y.C., Y.Y., M.L., W.C., Y.W. and X.L. substantively revised the manuscript during the writing process. L.C.X supervised the overall process. All authors read and approved the submitted version.

## Acknowledgements

We acknowledge GWAS Catalog and FinnGen study for providing the GWAS summary statistics data of HF and CAD, the Tabula Sapiens Single-Cell Dataset for providing the single-cell data, and the Drugbank for providing drug data, we are grateful to all the participants in these studies.

