## Additional file 2 for "Multi-trait genome-wide analysis identified novel risk loci and candidate drugs for heart failure"

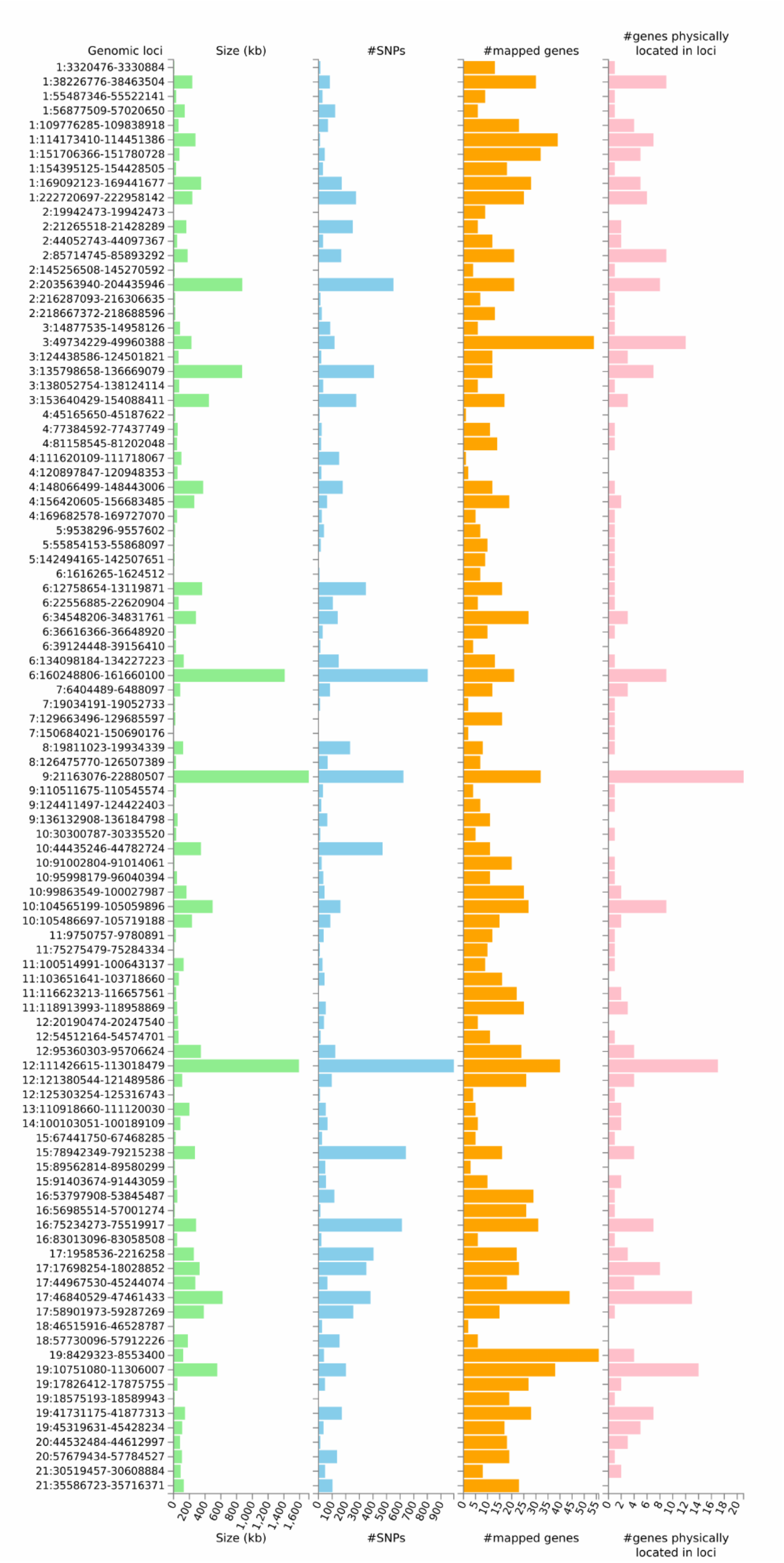


**Fig. S1 Summary of 99 HF risk loci.** Bar graphs depicting the size of the genomic locus (left one), number of candidate SNPs in the locus (left two), number of mapped genes in the genomic locus (right two) and the number of genes physically locating within the genomic locus (right one). Candidate SNPs are in LD (r2=0.1) of one of the independent significant SNPs.

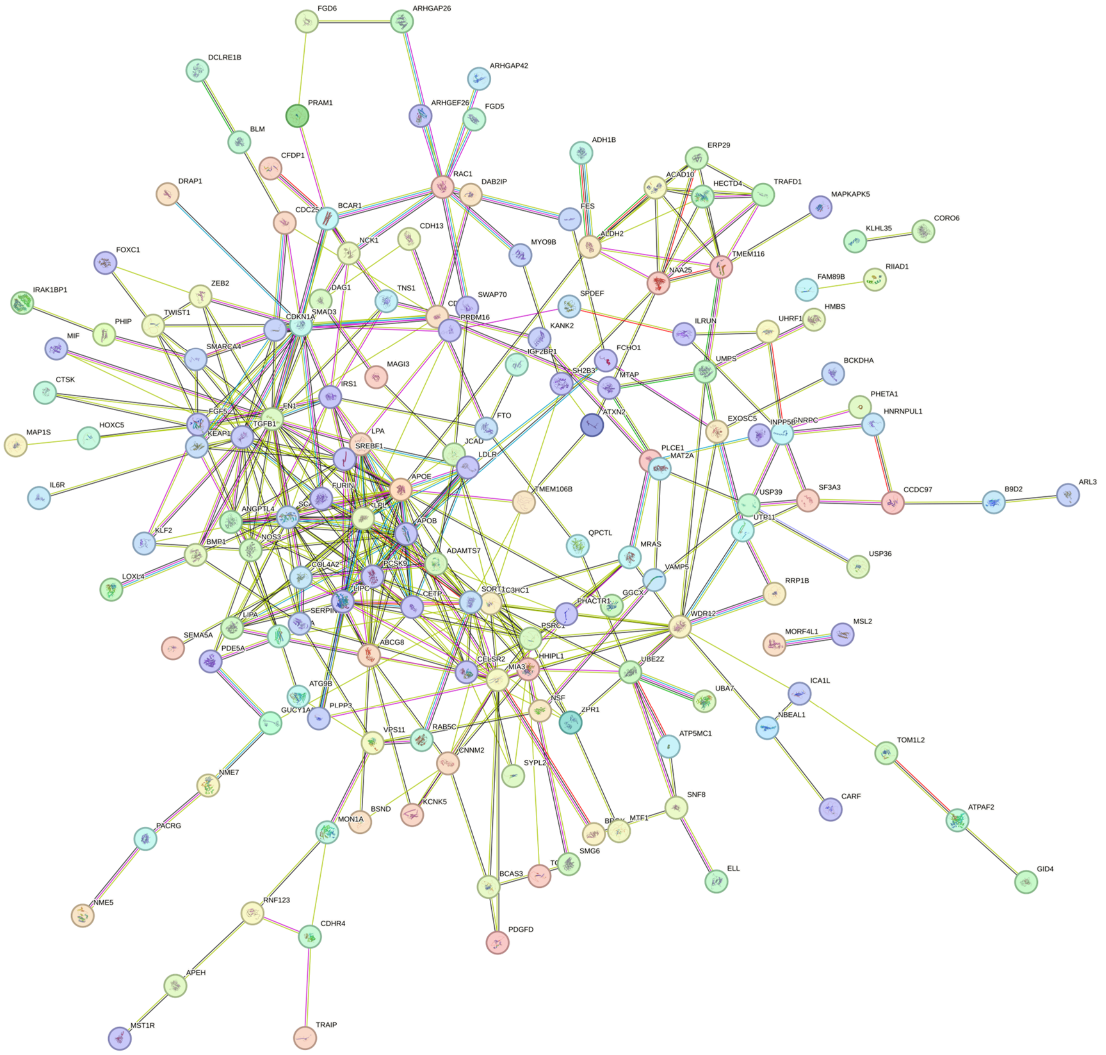


**Fig. S2 Protein–protein interaction network of 215 HF risk genes identified by transcriptome-wide association study (TWAS).** The reported network, including both direct (physical) and indirect (functional) associations, was based on the STRING v12.0 database. Only medium-confidence interactions between proteins are reported (interaction score > 0.4), while disconnected nodes in the network are hidden. Each node represents all the proteins produced by a single protein-coding gene locus, while edges represent protein–protein associations. Line color indicates the type of interaction evidence: light blue— from curated databases; purple—experimentally determined; green—gene neighborhood; red—gene fusions; blue—gene co-occurrence; yellow—text mining; black—co-expression; and violet—protein homology.
